# Dengue epidemic in a non-endemic zone of Bangladesh: clinical and laboratory profiles of patients

**DOI:** 10.1101/2020.06.08.20126094

**Authors:** Md. Abdur Rafi, Ashrafun Nahar Mousumi, Reejvi Ahmed, Md. Rezwanul Haque Chowdhury, Md. Abdul Wadood, Md. Golam Hossain

## Abstract

**Backgrounds:** Approximately, half of the population in the world including tropical and sub-tropical climates region is at risk of dengue. Being an endemic country, Bangladesh has experienced the largest dengue epidemic in 2019. The present study aimed at evaluating the clinical and laboratory profile of dengue patients in northern Bangladesh during the epidemic.

**Methods:** This cross-sectional study included 319 serologically confirmed dengue patients admitted in Shaheed Ziaur Rahman Medical College Hospital in Bogra district. It is one of the main tertiary care hospitals in northern Bangladesh. Data were collected from July to September 2019. Patients’ clinical and laboratory data were extracted from clinical records. Patients were classified into two classes according to the WHO 2009 dengue classification such as (i) non-severe dengue and (ii) severe dengue. Chi-square test and independent t-test were used in this study.

**Results:** Of the 319 patients, 94.1% had non-severe dengue and the remaining 5.9% had severe dengue (severe plasma leakage 68.4%, severe organ involvement 68.4%, and severe clinical bleeding 10.5%). Most of the patients were suffering from primary dengue infection. The most common clinical presentation was fever followed by headache and myalgia. Vomiting and abdominal pain were the most prevalent warning signs. The common hematological findings on admission were leukopenia (63.3%), thrombocytopenia (30.4%) and increased hematocrit (26.6%). Raised serum ALT or AST was observed in 14.1% cases whereas raised serum creatinine was observed in 6.6% cases. Signs of plasma leakage (pleural effusion, respiratory distress, and ascites, rise of hematocrit >20% during hospital stay) and hepatic or renal involvement (serum ALT >42UI/L or serum creatinine >1.2 mg/dL) on admission were mostly associated with severe dengue.

**Conclusion:** The study provides clinical evidence on presentation as well as hematological and biochemical profile of dengue patients in northern Bangladesh that should be implicated in effective patient management.

**Author summary:** Dengue has become a significant public health concern worldwide in recent years especially for the South-East Asian, sub-Saharan African and Latin American countries. Bangladesh has experienced a number of outbreaks of dengue, the largest one occurred in 2019. Management of dengue cases during an epidemic is a major challenge for a limited resource country like Bangladesh. To predict the risk of developing severe dengue a combined evaluation of early symptoms and laboratory test profiles is necessary. Despite the fact, there is a lack of evidence of clinical and laboratory parameters of dengue patients in Bangladesh. Authors wanted to highlight the clinical features, and hematological and biochemical profiles of dengue patients diagnosed in Bogra district, a non-endemic zone situated in northern Bangladesh. The authors reported that fever, headache, and myalgia were the commonest presenting complaints of dengue patients whereas vomiting and abdominal pain was the most prevalent warning signs. Severe dengue was associated mostly with plasma leakage rather than hemorrhage and the rise of hematocrit during hospital stay was a predictor of severe dengue. These findings will guide physicians for prompt therapeutic management of dengue infection in the study area.

## Introduction

Dengue is an arboviral disease caused by Dengue virus (DENV) that transmitted to humans by infected mosquitoes of the species *Aedes aegypti* and sometimes *Aedes albopictus* [1]. It has become a significant public health concern worldwide in recent years especially for the South-East Asian, sub-Saharan African and Latin American countries [2]. Almost half of the world’s population, living in the tropical and subtropical climate of these regions is at risk of dengue infection [3]. More than 390 million dengue virus infections occur worldwide every year causing more than 20,000 deaths [4, 5]. Bangladesh is situated in the dengue endemic zone of the South-East Asian region. The first dengue epidemic was reported here in 2000 with 5,521 confirmed dengue cases and 92 deaths [6]. Since then, Bangladesh has experienced a number of outbreaks of dengue. The largest one occurred in 2019, peaked from July to October affecting more than 100 thousand people and caused 164 deaths all over the country [7]. Rapid and unplanned urbanization without proper sanitation facilities contributing fertile breeding areas for mosquitoes, lack of vector control and climatic changes contributed to widespread dengue infection [8].

DENV, a single-stranded RNA virus of the genus Flavivirus, has four serotypes (DEN 1 to 4). Different serotypes can circulate simultaneously during an epidemic with a potential risk of infecting a person as many as four times, once with each serotype. Subsequent infections with different DENV serotypes increase disease severity [1]. Dengue infection can be clinically presented with varying severity ranging from asymptomatic infection or self-limiting influenza-like illness to a potentially fatal dengue hemorrhagic fever or dengue shock syndrome [9]. It is evidenced that all four serotypes were circulating in Bangladesh during different epidemics [10, 11] and during the current epidemic re-emergence of DEN-3 serotype was suspected, resulting in a more severe form of dengue infection [12].

Management of dengue cases during an epidemic is a major challenge for a limited resource country like Bangladesh. WHO introduced a new dengue case definition in 2009 for more convenient clinical management and it has set a rational approach for early prediction of severe dengue, triage, and management of patients based on clinical and laboratory parameters [13]. It is important to trace the cases that are more likely to develop severe dengue to prioritize their management. This can help in reducing admission and economic burden on health care facilities of low risk patients, who can be managed with less intensive follow up in outpatient settings. To predict the risk of developing severe dengue a combined evaluation of early symptoms and laboratory test profiles is necessary [14]. Despite the fact, there is a lack of evidence of clinical and laboratory parameters of dengue patients in Bangladesh. There are some studies mostly confined in Dhaka, and the central region of the country [15-17]. However, the epidemic affected the whole country and almost half of the cases were reported from outside of Dhaka city [7].

The present study aimed to highlight the clinical features, and hematological and biochemical profiles of dengue patients diagnosed in Bogra district, a non-endemic zone situated in northern Bangladesh.

## Methods

### Setting and participants

This prospective cross-sectional study was conducted in Shaheed Ziaur Rahman Medical College Hospital (SZMCH), a 1000-bed tertiary care teaching hospital situated in Bogra city from July to September, 2019. All the adult patients (aged more than 18 years) diagnosed as dengue by serological evidence (positive dengue NS1 antigen or IgM antibody) admitted to SZMCH according to the clinical judgment and decision of the physician on duty, and who met the inclusion criteria were included in the study. The first dengue case of the year was admitted to SZMC during the first week of June and continued to increase the number of cases up to last week of September. Last patient of our study was included on September 24, and we observed for two more weeks for any new case. As there was no new case during this period, the investigators decided to stop data collection. A total of 319 patients were included in the study.

### Inclusion and exclusion criteria

We followed two inclusion criteria such as (i) patients presented with clinical criteria of dengue fever, defined as a history of acute fever with at least two of the following symptoms: headache, retro-orbital or ocular pain, myalgia, arthralgia, rash, a positive tourniquet test (defined as the presence of ≥ 20 petechiae per 1 square inch), or leukopenia (defined as a white blood cell count 5000 cells/mm^3^); and (ii) a confirmed dengue viral infection, defined as positive tests of either specific dengue NS1 antigen if the blood sample was obtained on the sixth or later day of onset of symptoms, or IgM antibody if the blood sample was obtained on the sixth or later day of onset of symptoms using enzyme-linked immunosorbent assays (ELISA) from serum samples on admission. Patients with mixed infection (simultaneous infection by other organisms like *Plasmodium* or chikungunya virus along with dengue virus), or who were pregnant, were excluded from this study.

### Data collection procedure

Dengue related information including baseline characteristics, clinical parameters, and laboratory findings were recorded from patients in a pre-designed case-report form. Clinical examinations were performed by a physician on admission for each study participant. The NS1 antigen, IgM and IgG antibody detection were done by enzyme-linked immuno-sorbent assay (ELISA) method in the microbiology laboratory of SZMC for the diagnosis of dengue before admission of the patients. Other laboratory investigations including complete blood count (using Sysmex XN-2000™ Hematology Autoanalyzer, Sysmex Corporation), serum ALT, AST, and creatinine were performed on patient admission and complete blood count was repeated everyday to track hematocrit and platelet count changes. The severity of dengue was categorized on the date of discharge according to the WHO’s 2009 classification as non-severe and severe dengue [8]. Patients with non-severe dengue were subcategorized into two groups depending on the presence or absence of warning signs. All dengue patients in this study received standard care according to WHO guidelines [8].

### Dengue case definitions

Non-severe dengue without warning signs was defined as having acute fever with at least two of the following criteria: headache, myalgia, arthralgia, nausea, vomiting, rash, a positive tourniquet test, or leukopenia. Warning signs included: abdominal pain or tenderness, persistent vomiting (vomiting with signs of dehydration), clinical fluid accumulation, bleeding from mucosal areas including nose, gums, gastrointestinal tract or vagina, lethargy, restlessness, liver enlargement >2 cm, increase in hematocrit (≥20%) concurrent with rapid decrease in platelet count (≤100000/μL). Severe dengue was defined as having (i) severe plasma leakage, defined as plasma leakage with shock or respiratory distress (respiratory rate ≥24 breaths/min with oxygen saturation <95 % in room air and/or requiring oxygen therapy), (ii) severe clinical bleeding, defined as spontaneous bleeding from mucosal areas that necessitates a blood transfusion, or (iii) severe organ involvement, defined as AST >1000 IU/L and/or ALT >1000 IU/L, serum creatinine ≥2 times above baseline, myocarditis, and/or encephalitis [13]. Thrombocytopenia, leukopenia, raised hematocrit, raised ALT, raised AST and raised creatinine were defined as a platelet count of 100,000 platelets/μL or less, white blood cell count 5000 cells/μL, as >48% or ≥20% increase than baseline, as >42 IU/L, as >37 IU/L and as >1.2 mg/dl respectively [19, 20]. The dengue infection was defined as secondary if the IgM/IgG ratio was less than 1.2 [13].

### Statistical analysis

All statistical analyses were carried out using SPSS (IBM version 22.0). Clinical and laboratory parameters among different groups were compared with the chi-square test or Fisher’s exact tests, as appropriate in case of numerical variable and t-test or ANOVA, as appropriate in case of continuous variables. For the case of t-test and ANOVA, we checked the standard assumptions of t-test and ANOVA. All tests were interpreted with p < 0.05 indicating statistical significance.

### Ethical approval and consent of the participants

Ethical approval for this study was obtained from the Institutional Review Board of Shaheed Ziaur Rahman Medical College, Bogra, Bangladesh [Ref. No. SZMC/2019/132]. Before the commencement of the study, written consent was obtained from the participating patients after informing them of the objectives of the study and their right to remain or opt out of the study if they feel uncomfortable.

## Results

### Baseline characteristics of dengue cases

A total of 319 hospitalized patients with serologically confirmed dengue viral infection were included in this study. Among them, 70% were male and patients’ mean (SD) age was 33.0 (14.07) years, most of them were young adult (age less than 40 years). 304 (95.3%) patients were confirmed by positive dengue NS1 antigen and 15 (4.7%) patients were confirmed by positive dengue specific IgM antibody (Table 1). It was found that 291 (91.2%) and 28 (8.8%) patients had primary dengue and secondary dengue respectively. It was noted that the travel history to endemic zone was present in 186 (58.3%) patients. Most of the patients were admitted during the febrile phase of dengue. Among the 319 patients with confirmed dengue infection, 300 (94%) had non-severe dengue including dengue without warning signs (155, 48.6%) and dengue with warning signs (145, 45.5%). The remaining 19 patients (5.9%) were diagnosed as severe dengue. Among the patients with severe dengue, 13 (68.4%), 13 (68.4%) and 2 (10.5%) had severe plasma leakage, organ involvement, and clinical bleeding. Of the 13 patients, 11 were presented with severe plasma leakage, among them 1 (7.7%), 7 (53.8%) and 3 (23%) presented with shock, pleural effusion and ascites respectively. Patients with organ involvement mostly (6 (46.2%)) presented with liver involvement followed by kidney involvement (7 (53.8%)).

**Table 1:** Socio-demographic and diagnostic characters of dengue case

| Characteristics | Total | Severe dengue,<br>N (%) | Non-severe<br>dengue, N (%) | p-value |
| --- | --- | --- | --- | --- |
| Age (years), Mean (SD) | 33.00 (14.07) | 36.00 (16.62) | 32.81 (13.90) | 0.339 |
| Age category (years) |  |  |  |  |
| 18-30 | 155 | 11 (7.1) | 144 (92.9) | 0.262 |
| 31-40 | 90 | 4 (4.4) | 86 (95.6) |  |
| 41-50 | 36 | 0 (0.0) | 36 (100.0) |  |
| 51-60 | 25 | 2 (8.0) | 23 (92.0) |  |
| >60 | 13 | 2 (15.4) | 11 (84.6) |  |
| Sex |  |  |  |  |
| Female | 97 | 2 (2.1) | 95 (97.9) | 0.07 |
| Male | 222 | 17 (7.7) | 205 (92.3) |  |
| Residence |  |  |  |  |
| Rural | 172 | 13 (7.6) | 159 (92.4) | 0.191 |
| Urban | 147 | 6 (4.1) | 141 (95.9) |  |
| Family income |  |  |  |  |
| Higher | 13 | 0 (0.0) | 13 (100.0) | 0.173 |
| Lower | 116 | 4 (3.4) | 112 (96.6) |  |
| Middle | 190 | 15 (7.9) | 175 (92.1) |  |
| Travel history to endemic zone |  |  |  |  |
| Positive | 186 | 9 (4.8) | 177 (95.2) | 0.319 |
| Negative | 133 | 10 (7.5) | 123 (92.5) |  |
| Comorbidities (Yes) |  |  |  |  |
| Diabetes | 21 | 2 (9.5) | 19 (90.5) | 0.624 |
| Hypertension | 22 | 4 (18.2) | 18 (81.8) | 0.033 |
| Liver disease | 2 | 0 (0.0) | 2 (100.0) | 1.00 |
| Heart disease | 3 | 2 (66.7) | 1 (33.3) | 0.010 |
| Dengue NS1 antigen |  |  |  |  |
| Positive | 304 | 19 (6.2) | 285 (93.8) | 0.612 |
| Negative | 15 | 0 (0.0) | 15 (100.0) |  |
| IgM |  |  |  |  |
| Positive | 56 | 2 (3.6) | 54 (96.4) | 0.545 |
| Negative | 263 | 17 (6.5) | 246 (93.5) |  |
| Type of infection |  |  |  |  |
| Primary | 291 | 13 (4.5) | 278 (95.5) | 0.000 |
| Secondary | 28 | 6 (21.4) | 22 (78.6) |  |
| Phase on admission |  |  |  |  |
| Febrile | 213 | 8 (3.8) | 205 | 0.002 |
| Critical | 77 | 11 (14.3) | 66 |  |
| Recovery | 29 | 0 (0.0) | 29 |  |

### Clinical presentations of dengue cases

The most common clinical feature on presentation was fever (92.5%) followed by headache (72.7%) and myalgia (71.5%). Vomiting was the most common warning sign (34.2%) followed by abdominal pain 95 (29.8%). Petechiae or rash was the most common evidence of bleeding (15.7%) followed by melena (11.9%), sub-conjunctival hemorrhage (8.8%) and gum bleeding (7.8%). The majority of symptoms at admission were similar between patients with severe and non-severe dengue with some exceptions, which were more frequently associated with severe dengue. These were not having fever, abdominal pain, respiratory distress, hepatomegaly (> 2cm), ascites, pleural effusion, peripheral edema and shock (Table 2).

**Table 2:** Clinical presentations of dengue cases at the time of hospital visit (n=319)

| Clinical feature (Yes) | Total | Severe dengue, N (%) | Non-severe dengue, N (%) | p-value |
| --- | --- | --- | --- | --- |
| Fever | 295 | 15 (5.1) | 280 (94.9) | 0.044 |
| Myalgia | 228 | 17 (7.5) | 211 (92.5) | 0.113 |
| Headache | 232 | 10 (4.3) | 222 (96.7) | 0.043 |
| Retro-orbital pain | 150 | 3 (2.0) | 147 (98.0) | 0.008 |
| Abdominal pain | 95 | 12 (12.6) | 83 (87.4) | 0.001 |
| Vomiting | 109 | 8 (7.3) | 101 (92.7) | 0.452 |
| Diarrhea | 138 | 7 (5.1) | 131 (94.9) | 0.560 |
| Constipation | 12 | 0 (0.0) | 12 (100.0) | 0.627 |
| Respiratory distress | 7 | 7 (100.0) | 0 (0.0) | 0.000 |
| Hepatomegaly | 23 | 9 (39.1) | 14 (60.9) | 0.000 |
| Cough | 9 | 2 (22.2) | 7 (77.8) | 0.094 |
| Bleeding manifestation |  |  |  |  |
| Melena | 38 | 1 (2.6) | 37 (97.4) | 0.490 |
| Hematemesis | 10 | 0 (0.0) | 10 (100) | 0.650 |
| Rash | 50 | 2 (4.0) | 48 (96.0) | 0.748 |
| Gum bleeding | 25 | 0 (0.0) | 25 (100.0) | 0.380 |
| Epistaxis | 8 | 0 (0.0) | 8 (100.0) | 1.000 |
| Hematuria | 9 | 1 (11.1) | 8 (88.9) | 1.000 |
| Sub-conjunctival<br>hemorrhage | 28 | 0 (0.0) | 28 (100.0) | 0.239 |
| Other bleeding manifestation (e.g. menorrhagia, joint bleeding etc.) | 10 | 3 (30.0) | 7 (70) | 0.017 |
| Features of plasma leakage |  |  |  |  |
| Ascites | 3 | 3 (100.0) | 0 (0.0) | 0.000 |
| Pleural effusion | 7 | 7 (100.0) | 0 (0.0) | 0.000 |
| Edema | 5 | 3 (60.0) | 2 (40.0) | 0.002 |

\* $\chi^2$ -test done to compare groups according to presence (Yes) or absence (No) of a specific clinical feature

### Hematological and biochemical parameters of dengue cases

It was observed that the most common hematological finding on admission was leukopenia (< 5000) (63.3%) and thrombocytopenia (< 100 000) (30.4%). Hematocrit > 48% were noted in 85 (26.6%) during total hospital stay whereas >20% increase in hematocrit was observed in 74 (23.2%) patients. Alanine aminotransferase (ALT) level >42 IU/L and aspartate aminotransferase (AST) level >37 IU/L were observed in 40 (12.5%) cases and 16 (5.0%) cases respectively. Serum creatinine level >1.2 mg/dl was observed in 21 (6.6%) cases (Table 3). Most laboratory results among patients with severe and non-severe dengue were similar with the exceptions of rise of hematocrit, ALT and serum creatinine which were significantly higher among patients with severe dengue compared to those with non-severe dengue.

**Table 3:** Hematological and biochemical parameters of dengue cases (n = 319)

| Parameters | Total (n = 319) | Severe dengue (n = 19) | Non-severe dengue (n = 300) | p-value |
| --- | --- | --- | --- | --- |
| Hematological parameters |  |  |  |  |
| HCT (%), Mean (SD) | 39.97 (6.23) | 39.68 (8.56) | 39.99 (6.07) | 0.309 |
| Leucocyte count (cells$\times 10^3$ cells/μL), Mean (SD) | 5.4 (3.1) | 6.0 (6.1) | 5.4 (2.8) | 0.634 |
| Platelet count (cells$\times 10^3$/μL), Mean (SD) | 138 (82.4) | 110 (49.4) | 139.8 (83.8) | 0.127 |
| Categorical data |  |  |  |  |
| HCT (%) |  |  |  |  |
| $\leq 48$ | 292 | 19 (6.5) | 273 (93.5) | 0.242 |
| $> 48$ | 27 | 0 (0.0) | 27 (100.0) | |
| Rise of HCT (during hospital stay) |  |  |  |  |
| $\leq 20\%$ | 245 | 10 (4.1) | 235 (95.9) | 0.010 |
| $> 20\%$ | 74 | 9 (12.2) | 65 (87.8) | |
| Leucocyte count (cells/μL) | | | | |
| $\leq 5000$ | 202 | 14 (6.9) | 188 (93.1) | 0.334 |
| $> 5000$ | 117 | 5 (4.3) | 112 (95.7) | |
| Platelet count (cells/μL) | | | | |
| $\leq 100\ 000$ | 97 | 5 (5.2) | 92 (94.8) | 0.689 |
| $> 100\ 000$ | 222 | 14 (6.3) | 208 (93.7) | |
| Biochemical parameters |  |  |  |  |
| Serum ALT (IU/L) | 248.69<br>(367.45) | 676.73 (557.68) | 130.98 (165.73) | 0.009 |
| Serum AST (IU/L) | 267.50<br>(396.42) | 612.50 (561.61) | 95.00 (45.97) | 0.074 |
| Serum Creatinine (mg/dL) | 1.34 (0.75) | 2.6885 (0.82) | 1.04 (0.23) | 0.000 |
| Categorical data |  |  |  |  |
| Serum ALT (IU/L) |  |  |  |  |
| $> 42$ | 40 | 11 (27.5) | 29 (72.5) | 0.000 |
| $\leq 42$ | 279 (87.5) | 8 (2.9) | 271 (97.1) | |
| Serum AST (IU/L) |  |  |  |  |
| $> 37$ | 16 (5.0) | 6 (37.5) | 10 (62.5) | 0.000 |
| $\leq 37$ | 303 (95.0) | 13 (4.3) | 290 (95.7) | |
| Serum creatinine (mg/dL) |  |  |  |  |
| $> 1.2$ | 21 (6.6) | 12 (57.1) | 9 (42.9) | 0.000 |
| ≤ 1.2 | 298 (93.4) | 7 (2.3) | 291 (97.7) |  |

## Discussion

The dengue epidemic has become a major public health threat for many South-East Asian endemic countries. A number of outbreaks of dengue occurred in Bangladesh, mostly affecting Dhaka, the capital city and the central region. Though the exact cause of being Dhaka an endemic zone of dengue in Bangladesh, it is likely that the humid weather and more source of mosquito breeding (stagnant clean water during rainy season) due to unplanned urbanization and poor drainage system could make the city vulnerable for dengue infection. A nationwide sero-prevalence study reported much lower prevalence in the northern region of the country compared to the central region and most of these cases were suspected to be referred from endemic zones [21]. Despite the fact, during the 2019 epidemic a huge number of cases were reported from this region [7] and travel history to endemic zone was not a significant risk factor of dengue infection of these patients, as our study found. It is a matter of concern that dengue is not confined to the reported endemic zone of the country; rather it has become a national health emergency.

Appropriate clinical management of the huge number of patients during an epidemic is a great challenge for a poor resource country like Bangladesh. To address the issue WHO revised the previous classification system and introduced a newer version in 2009 with an intention of more convenient management based on clinical and laboratory parameters. Despite the fact, the implication of this classification resulted in a higher number of hospital admissions of dengue fever increasing the burden on vulnerable health care system [22, 23]. It is important to identify the patients who are potentially at risk of developing severe disease and need more intensive monitoring by clinical and primary laboratory evaluation. It was necessary to explore these parameters in the context of Bangladeshi patients as there is hardly any evidence of clinical, hematological and biochemical profile of dengue patients based on the updated case definition. The present study was aimed at meeting the knowledge gap which would help to improve the predictive and diagnostic process of dengue.

Clinical manifestations and laboratory profiles of our dengue patient cohort were quite similar to the patients of previous dengue epidemics in Bangladesh [15, 16, 24, 25]. Fever, headache, and myalgia were the most common clinical presentations of our patients. In hematological evaluation, leukopenia, thrombocytopenia, and raised hematocrit level were the most frequent findings. Petechiae or skin rash was commonest bleeding manifestation among the present cohort as evidenced in previous studies conducted in the endemic area [15, 24], though the prevalence was much lower here. However, severe dengue was mainly manifested as severe plasma leakage and severe organ involvement in our study population rather than dengue hemorrhagic fever, which was more prevalent in previous epidemics [24, 25]. Severe plasma leakage was also a more common manifestation of severe dengue during a recent epidemic in Singapore [26].

Most of the patients of the present study were suffering from primary dengue while the ratio of primary and secondary dengue was almost equal in previous epidemics that occurred in Dhaka city [24]. The sero-prevalence of dengue was reported as much lower in our study region compared to Dhaka city (21). Secondary infection was more likely to be associated with severe dengue among our study population, consistent with findings of other studies [18, 27, 28].

Fever, headache, retro-orbital pain and other constitutional symptoms were associated with non-sever dengue. The basic pathophysiology of this finding is that these non-specific symptoms of viral febrile illness are mostly found in the febrile phase of dengue, whereas severe dengue is usually manifested during the critical phase, when there is increased risk of plasma leakage and shock [13, 29]. For this phenomenon, patients, who were admitted during the critical phase of the disease course developed severe dengue more frequently according to our finding, as evidenced in a previous study too [29]. Warning signs like abdominal pain and hepatomegaly were associated with severe dengue among our patients. Abdominal pain along with persistent vomiting was reported as strong predictors of severe dengue by many researchers [18, 26, 30-32]. Pleural effusion along with respiratory distress and ascites were the most common presentations of severe plasma leakage, consistent with a recent study conducted in Singapore [26]. Ascites was not only an important predictor of dengue shock syndrome [14, 26] but also an early predictor for admission to the intensive care unit [33].

Among hematological parameters, leukopenia and thrombocytopenia were the most common findings on admission though there was no significant difference in these parameters between severe and non-severe dengue patients. However, rise in hematocrit more than 20% during hospital stay was associated with severe dengue in our patient cohort, which is similar to previous findings, though thrombocytopenia was also reported as a predictor of severe dengue in these studies [14, 30, 34]. Raised hematocrit is the evidence of plasma leakage whereas thrombocytopenia may cause severe hemorrhage, the two main varieties of severe dengue [13]. Most of the patients’ suffering from severe dengue of our study cohort presented as severe plasma leakage rather than severe hemorrhage, so thrombocytopenia was not significantly associated with severe dengue. Our result suggests that severe organ involvement can be predicted from early rise of biomarkers, especially serum ALT in case of liver involvement and serum creatinine in case of renal involvement. Similar results were reported by others [18, 26, 35-37].

According to our findings, almost half of the admitted dengue patients in SZMCH had no warning sign, which had made them vulnerable for developing severe dengue, as defined by the WHO. There is a justifiable space for argument whether admitting these patients was appropriate or not. As our study and previous ones suggested, warning signs like abdominal pain, hepatomegaly as well as persistent vomiting were more likely associated with developing severe disease, these clinical features should be taken into account when screening a dengue patient for admission, especially in resource-poor settings like Bangladesh. Moreover, increasing hematocrit >20% during hospital stay, raised liver enzyme and serum creatinine beyond upper margin of normal value were associated with severe dengue. So, screening patients with these laboratory parameters on first visit with periodic follow up keeping them on outdoor management may reduce the hospital burden of those admitted patients for whom outdoor management was potentially enough.

Although the present study was the first attempt to report dengue patient profile from a non-endemic region of the country, it has some limitations: (i) patients were recruited from a specific geographic territory, which is not in the dengue-endemic zone of the country. So our data may not be representative of dengue patients of the whole country; (ii) only hospitalized patients in the study center (SZMCH) were included in this study and as a result, a number of non-severe dengue patients, who were managed in outpatient settings and those who were treated in other facilities as well as did not visit hospital due to mildness of symptoms or other causes were missed, so the patients included in this study do not represent all the patients with dengue in this region; (iii) we confirmed dengue based on serological test, RT-PCR to detect viral RNA and serotyping of DENV was not done; (iv) hydration and volume expansion could have had an effect on hematocrit that could potentially bias the data. Regardless of the depicted limitations, the present study ultimately provides the baseline data on clinical, hematological and biochemical profiles of dengue patients of the respective region of the country as well as generate evidence of clinical and laboratory parameters to predict the development of severe dengue.

## Conclusions

Management and prognosis of dengue are mostly based on clinical presentations as well as laboratory findings like hematological and biochemical parameters. The present study reported that fever, headache, and myalgia were the commonest presenting complaints of dengue patients whereas vomiting and abdominal pain was the most prevalent warning signs. Severe dengue was associated mostly with plasma leakage rather than hemorrhage and the rise of hematocrit during hospital stay was a predictor of severe dengue. We hope and believe that these findings will guide physicians for prompt therapeutic management of dengue infection in the study area.

## Data Availability

Patient level data will be available on request, provided that an approval is given from the Ethical Review Committee of Shaheed Ziaur Rahman Medical College, Bogra, Bangladesh.

## List of abbreviations

DENV: Dengue Virus
ALT: Alanine aminotransferase
AST: Aspartate aminotransferase
ELISA: Enzyme-Linked Immunosorbent Assay
NS1: Non-specific antigen-1
IgM: Immunoglobulin M
IgG: Immunoglobulin G
SZMCH: Shaheed Ziaur Rahman Medical College Hospital
WHO: World Health Organization

## Consent for publication

Not applicable.

## Author Contributions

Conceptualization: Md. Abdur Rafi, Ashrafun Nahar. Formal analysis: Md. Abdur Rafi, Md. Abdul Wadood, Md. Golam Hossain. Investigation: Md. Abdur Rafi, Ashrafun Nahar, Reejvi Ahmed, Md. Rezwanul Haque Chowdhury. Methodology: Md. Abdur Rafi, Ashrafun Nahar. Resources: Md. Abdur Rafi. Supervision: Md. Golam Hossain. Writing – original draft: Md. Abdur Rafi, Md. Golam Hossain. Writing – review & editing: Md. Abdur Rafi, Ashrafun Nahar, Reejvi Ahmed, Md. Rezwanul Haque Chowdhury, Md. Abdul Wadood, Md. Golam Hossain.

## Acknowledgements

The authors would like to acknowledge Fazle Rabbi Chowdhury, department of Medicine, Bangabandhu Sheikh Mujib Medical University, Dhaka, Bangladesh, Md. Zakir Hossain and Md. Fakhrul Islam, department of medicine, Shaheed Ziaur Rahman Medical College, Bogura, Bangladesh for their contribution in conceptualization of the study and reviewing the manuscript. They would also like to thank Shahajada Farhan, Mahbuba Afrin, Md. Ashikur Rahman, Md. Abdulla Al Faraby, Plaban Chandra Sarker and Md. Nahid Hasan for their support in data collection. The authors would also like to express their sincere gratitude to all study participants and the staff engaged in the study.

